# *“We tend to get pad happy”*: a qualitative study of health practitioners’ perspectives on continence care for older people in hospital

**DOI:** 10.1101/2020.12.05.20234690

**Authors:** John Percival, Katharine Abbott, Theresa Allain, Rachel Bradley, Fiona Cramp, Jenny Donovan, Candy McCabe, Kyra Neubauer, Sabi Redwood, Nikki Cotterill

## Abstract

**Background:** Bladder and bowel control difficulties affect twenty and ten per cent of the UK population respectively, touch all age groups and are particularly prevalent in the older (65+) population. However, the quality of continence care is often poor, compromising patient health and wellbeing, increasing the risk of infection and is a predisposing factor to nursing and residential home placement.

**Objective:** To identify factors that help or hinder good continence care in hospital.

**Methods:** We conducted 27 qualitative interviews with nursing, medical and allied health practitioners in three hospitals. We used a purposive sample and analysed data thematically, both manually and with the aid of NVivo software.

**Results:** Interviews revealed perspectives on practice promoting or inhibiting good quality continence care, as well as suggestions for improvements. Good continence care was said to be advanced through person-centred care, robust assessment and monitoring, and a proactive approach to encouraging patient independence. Barriers to quality care centred on lack of oversight, automatic use of incontinence products and staffing pressures. Suggested improvements centred on participatory care, open communication and care planning with a higher bladder and bowel health profile. In order to drive such improvements, hospital-based practitioners indicate a need and desire for regular continence care training.

**Conclusions:** Findings help explain the persistence of barriers to providing good quality care for patients with incontinence. Resolute continence promotion, in hospitals and throughout the NHS, would reduce reliance on products and the accompanying risks of patient dependency and catheter associated gram negative bacteraemia. Robust assessment and care planning, open communication and regular continence care training would assist such promotion and also help mitigate resource limitations by developing safer, time-efficient continence care.

## INTRODUCTION

Continence care is a fundamental aspect of health service provision and has a substantial impact on individuals and the NHS.[1-4] Over 14 million adults in the UK experience bladder control problems and 6.5 million have bowel control difficulties.[5] Incontinence, defined as any involuntary loss of urine or the inability to control the bowels,[6] disproportionately affects older people, although older age is not a determining factor.[7,8] Incontinence is more common than heart disease, breast cancer or diabetes among older women, yet its public health profile is undeveloped, demoting its importance and quality assurance safeguards.[9,10]

Incontinence brings significant personal and public costs. Quality of life can be reduced by isolation, anxiety, loneliness and depression, with self-esteem undermined by decreased independence, mobility and health.[11-15] Furthermore, older people are commonly too embarrassed to seek help and may reduce food and fluid intake, along with social contact and activity, so as to mitigate continence ‘accidents’.[7,16-18] Annual continence care costs to the NHS have risen from £77m in 2006/7 to over £200m in 2010/11.[19,20], with more recent data difficult to ascertain. Catheter associated urinary tract infections (CAUTI) are estimated to cost the NHS up to £99m per year, while pressure ulcers, often caused by poor care associated with incontinence, cost the NHS between £1.4 and £2.1 billion annually.[21] Additionally, inadequate management of incontinence can escalate costs resulting from morbidity, unnecessary and sometimes lengthy hospitalisation, and raised mortality rates.[20,21]

The quality of continence care has been under scrutiny in recent years, with relevant guidance advocating early assessment and identification of symptoms, together with initial conservative management and individualised care.[22-28] Treatment of incontinence, however, has been found to be inadequate,[29,30] particularly for older adults.[31,32] Falls, pressure ulcers and moisture lesions are directly linked to poor continence care in older adults.[33,34] National audits and guidance regarding continence care have identified that healthcare professionals in acute care settings inconsistently identify, assess, manage and treat incontinence. [35-37] The Francis public inquiry report referred to continence as ‘this most basic of needs’ and yet it was the area of care most frequently singled out for complaint.[38] Reports have also raised concern in respect of insufficient staff support and education, weak service integration and liaison, inconsistent care planning and poor communication regarding continence care.[29,37]

An over-reliance on certain products, such as incontinence pads and catheters, has also compromised the quality of continence care. Between 17 and 25 per cent of patients in acute care have an indwelling urinary catheter (IUC), of which at least a quarter are unnecessary.[39] Moreover, sixty per cent of urinary tract infections relate to catheter use and almost a third of medical and surgical inpatients are inappropriately treated with catheters.[20,39] Incontinence pads are sometimes not the right type or size, which can affect patient hygiene, skin integrity and dignity.[17] Nurses often rely on containment methods, such as pads, indwelling catheters and penile sheaths, rather than help patients maintain continence.[9,40] Training programmes on continence care are available but these are delivered in less than half of acute hospitals,[26,29,31,41] perhaps because of the low priority placed on this fundamental aspect of care.[9,35] Additionally, staff requiring help or advice in respect of continence care practice are unlikely to have access to local specialist nurses or services,[29,35] despite such provision having been strongly advocated in recent years.[20]

There is a clear need for improved continence care in the NHS [33,42] together with a better understanding of healthcare workers’ perspectives, as evidence is limited in this area.[31,33,38,42]. The aim of our study was to address these points by conducting in-depth interviews with nurses, doctors and allied health practitioners, in order to learn details of factors that help or hinder appropriate continence care for older adults in hospitals and perceived improvements to optimise such care.

## METHODOLOGY

### Study design and participant recruitment

A qualitative design was adopted, using semi-structured interviews with hospital based nursing, medical and allied health professional staff, of varying grades, to explore perspectives on the obstacles and challenges to evidence-based continence care and ways to optimise such care. The interview schedule was informed by previous audit data and existing evidence.[43] The main inclusion criterion was that research participants have responsibility for patients aged 65 and over. Recruitment was carried out through purposive sampling of staff groups to achieve representation across professional roles. Research participants were recruited in acute inpatient wards (mostly, though not exclusively, care of the elderly wards) at three hospitals in the South West of England. These sites were selected to represent two large, city teaching hospitals, (one with specific responsibility for Urology services) and one was a smaller, non-teaching centre. The lead researcher (JP) met with ward managers at each hospital to explain the research and provide participant information materials for display and distribution among staff. Representation from each staff group, nursing, medical and allied health, was achieved and the sample size determined during ongoing analysis and interpretation of emerging themes to ensure data saturation to achieve the study’s aims. Interviews took place in hospital and were carried out by JP, in accordance with research governance ethics protocols and with HRA approval (UWE REC reference: HAS.19.07.221).

### Data analysis

Interviews were audio recorded, transcribed verbatim and subject to thematic and content analysis.[44,45] Initial analysis involved close reading by JP of each transcript, at least twice, providing an opportunity for development of coding strategies in relation to emerging subject domains, themes and sub themes. Transcript data were then imported into the qualitative data management software package NVivo 12, with coding further refined by JP and the chief investigator (NC) using a coding frame devised to highlight theme connectivity and relevance to the primary areas of investigation.

## RESULTS

A total of 27 participants was evenly spread across the three hospital sites, as shown in Table 1. This total included 17 nursing staff, 5 allied health care professionals, 3 medical staff and 2 healthcare assistants.

**Table 1.** Hospital health care practitioner characteristics

| Prof Title | Age band | Gender | Practice years | Band | Ethnicity |
| --- | --- | --- | --- | --- | --- |
| Staff Nurse | 21-25 | F | 1.5 | 5 | Black British |
| Staff Nurse | 21-25 | F | <1 | 5 | White & Black Caribbean |
| Ward Sister | 26-30 | F | 6 | 7 | White British |
| Senior Staff Nurse | 26-30 | F | 6 | 6 | White British |
| Senior Staff Nurse | 51-55 | F | 9 | 6 | White British |
| Senior Staff Nurse | 26-30 | F | 5 | 6 | White British |
| Apprentice Healthcare Support Worker | 18-20 | F | <1 | 2 | White British |
| Therapy Technician | 31-35 | F | 6.5 | 4 | White British |
| Senior Nursing Assistant | 46-50 | F | 13 | 3 | White British |
| Staff Nurse | 56-60 | M | 29 | 5 | White British |
| Staff Nurse | 56-60 | F | 40 | 5 | White British |
| Student Nurse | 31-35 | F | <1 | N/A | White British |
| Physiotherapist | 22-25 | F | 1.5 | 5 | White British |
| Therapy Technician | 46-50 | F | 6 | 3 | White British |
| Consultant Physician | 36-40 | M | 14 | N/A | Indian |
| Nursing Assistant | 36-40 | F | 2 | 2 | White British |
| Staff Nurse | 46-50 | F | 2 | 5 | White British |
| Junior Sister | 31-35 | F | 9 | 6 | White British |
| Sister | 21-25 | F | 4 | 6 | White British |
| Assistant Nursing Practitioner | 31-35 | F | 5 | 4 | Black South African |
| Staff Nurse | 26-30 | F | 1 | 5 | White British |
| Consultant Geriatrician | 36-40 | M | 17 | N/A | White British |
| Physiotherapist | 41-45 | F | 15 | 6 | White British |
| Dementia Specialist Practitioner | 51-55 | F | 35 | 7 | White British |
| Occupational Therapist | 41-45 | F | 11 | 6 | White British |
| Healthcare Assistant | 51-55 | F | 3 | 3 | White British |
| Junior Doctor | 21-25 | F | <1 | F1 | White British |

The data reflected only slight differences in emphasis according to professional role, with no markedly divergent opinions arising. Key themes that emerged are therefore representative of all participants’ perspectives and concerns. These themes are: practice promoting good quality continence care; obstacles to good practice, and suggestions for improvements.

In order to safeguard confidentiality, the source of each participant interview excerpt included in this paper is identified using the practitioner’s study identification number.

### Practice promoting good quality continence care

Participants gave accounts of the constructive ways in which they addressed patients’ continence care needs, especially through: assessment and monitoring; planning ahead; person-centred care, and encouragement of independence.

#### Assessment and monitoring

According to interviewees, pro-forma hospital admission assessment procedures insufficiently captured continence care needs, due to *“variable”* or sometimes *“incomplete”* detail.

Further assessment and monitoring of skin condition, temperature and urine/faecal output therefore enabled staff to clarify a patient’s continence needs. Routine investigation also helped practitioners establish, for example, when incontinence pads could be dispensed with in favour of regular toileting, or whether, and under what circumstances, their incontinence would be manageable on return home. Moreover, the need for, and potential problems of, catheter use were said to be elucidated by regular monitoring.

#### Planning ahead

Participants told us the hospital environment could inhibit patients talking about their incontinence because they were embarrassed and feared drawing attention to themselves in this semi-public space. In this context, interviewees advocated the regular prompting of patients to use the toilet so they did not have to ask. Planning ahead was also important to a physiotherapist [B44], who commented that in the course of her work with catheterised patients who had restricted mobility, she strived to get a trial without catheter (TWOC) procedure (commonly referred to as ‘TWOC’d’) undertaken *“early”*, as this could help motivate patients to increase mobility as well as offset the risk of patients returning home still catheterised.

#### Person-centred care

Holistic, person-centred continence care was important to a number of participants, who spoke of getting to know and understand patients’ behaviour, body language and personal situation, in order to strengthen patient morale, dignity and self-confidence. Examples include a staff nurse [B42], who told of her work with catheterised patients with dementia whose psychological need to use the toilet was respected, and a nursing assistant [C22], who interpreted a patient’s change in behaviour as a *“communication tool”* alerting her to his urine retention. Underpinning these approaches to continence care was a commitment to building a rapport with patients. In this context, participants mentioned the relevance of sensitive and supportive communication, detailing how they would convey reassurance using a *“kind and caring face”* and act as an *“advocate”* for patients who had difficulty articulating their needs.

#### Encouragement of Independence

On all hospital wards included in the study, the ambition to help patients with incontinence *“get back”* to their original *“baseline”* was commonly reported by participants to be a primary objective in care-giving. Indeed, interviewees often conveyed the fundamental nature of this focus in their opening remarks about incontinence, with statements such as: *“We strive to get people as independent as possible”* [C23, staff nurse] and *“that’s what incontinence means to me, it’s trying to preserve our patients’ dignity and their independence”* [A09, senior staff nurse]. Chief amongst the ways in which participants said they helped patients with incontinence maximise independence and control was to work with them on their mobility difficulties so as to transfer more readily to the toilet. Interviewees indicated that assisting patients to mobilise in order to use the toilet was motivational, physically strengthening and positively impacted patients’ mental health, attributes contributing to self-worth and self-determination. In addition, such practice was said to be time-efficient:

> *If their continence improves by emptying their bladder and bowel [on the toilet rather than commode] and they feel a lot better because they have sat up [in privacy] in a good position, then they are going to have less interventions for their incontinence*. [C25, physiotherapist].

### Obstacles to good practice

It was clear from interviews that hospital practitioners faced certain barriers to providing good continence care, in particular: lack of oversight; over-reliance on incontinence products, and staffing pressures.

#### Lack of oversight

Participants pointed out that information gained from monitoring patients’ continence care was only useful if accurately and routinely recorded and, as this was not always the case, such information could not be relied upon:

> *This morning a lady was TWOC’d*..*she is mobile, gone to the toilet a couple of times last night but nobody has documented to see if she has passed urine or not or how much. [A04; senior nursing assistant]*

Interviewees also speculated that this laxity arose because of a tendency not to notice, or take stock of, the commonplace:

> *Continence care is an important area and the only thing I find is because it’s such basic care, it gets overlooked and forgotten about at times… some [staff] think there is nothing to worry about. But I think they should do. [B41, staff nurse]*

A ward sister concurred that continence care is *“forgotten about”*, claiming because other procedures and tasks are seen as more important, with the result that patients are *“kept in pads”* by unquestioning staff or are sent home with catheters that are not necessary. A dilatory attitude could also preclude consideration of proactive continence care strategies. In this respect a dementia specialist practitioner described how when she advised staff to engage in regular two hour toileting routines, to keep patients dry and stop them becoming agitated, she was sometimes viewed *“as though I have got snakes coming out of my head. It’s an old fashioned concept but it works”* [C26].

#### Over-reliance on incontinence products

The use of pads, without due regard to the possible health implications, led one nursing assistant to worry that this could *“encourage incontinence and becomes an obstacle to keeping people continent”* [B47]. A ward sister echoed this point when she drew the conclusion, *“We cause incontinence a bit”* [C21]. In addition to the question of staff inattention, discussed above, participants suggested a number of intrinsic reasons for over-reliance on products: custom and practice; expediency; and patient acceptance.

Custom and practice in respect of pad over-reliance was implied by many interviewees when speaking of pads being provided as a matter of course, without first determining need, suitability, benefit or risk. This convention was summed up by one staff nurse when admitting, *“we tend to get pad happy”* [C23]. A nursing assistant added weight to this concern by pointing out that *“registered nurses should prescribe continence products [but] it’s very rare that I have seen that done”* [B47], leaving her and her peers to make these clinical decisions.

Expediency, given staffing levels and pressures on time, was another reason for over-reliance on pads. A physiotherapist spoke of nurse colleagues being *“always up against the clock [so] it’s easier just to stick a pad on someone [and] not allow them to walk to the toilet”* [C25]. Although expediency was most often talked about in relation to over-reliance on pads, it was also said to contribute to overuse of catheters. Unnecessary provision of catheters was said to occur as a service convenience or because staff were too busy to review their use. A junior doctor noted that over-reliance on catheters and delay in their removal *“can be a very common situation” [C29]*.

Patients themselves were said to be, on occasion, complicit in the routine use of products such as incontinence pads, by accepting pads in order to reduce the burden on obviously over-stretched staff. As one staff nurse put it, *“they don’t want to bother the nurse”*. Another reason for patients’ assent in the regular use of pads was said to be anxiety and embarrassment about their incontinence, which could lead to dependency on pads as a method of masking their incontinence in a semi-public environment. An important consequence of product over-reliance was reported to be the weakening of patient independence and fostering of dependency.

#### Staffing pressures

A senior staff nurse [A01] referred to the benefit of having secured an extra nursing assistant to help patients with toileting but added that with the heavy workload on the ward, *“we are still struggling”*. The general pressure that hospital staff were under led one consultant geriatrician to reflect how patients’ incontinence needs could be missed if they were not highlighted by the patient, adding *“the pressures on hospital are to treat the primary condition and get them out… we don’t pick up [continence needs] as much as we could* do *because it’s not the thing that’s flagged up in front of you”* [C24]. Other staff agreed that key indicators of continence need could easily be overlooked, with one staff nurse saying by way of example, *“this patient might not have wee’d for an hour or two and urine output might have tailed off [which] might be missed”* [A06].

These concerns were often reinforced by reference to low staff to patient ratios, especially problematic on wards where many patients were bedbound or recovering from surgery, requiring two members of staff to support them with their needs.

### Suggested Improvements

Participants’ ideas on improvements to continence care practice coalesced around four issues: participatory care; open communication; robust planning, and staff training. These issues are represented in Figure 1, which depicts staff training as central to the other desired improvements and also to two (flagged) likely outcomes, emphasised throughout interviews.

**Figure 1.**
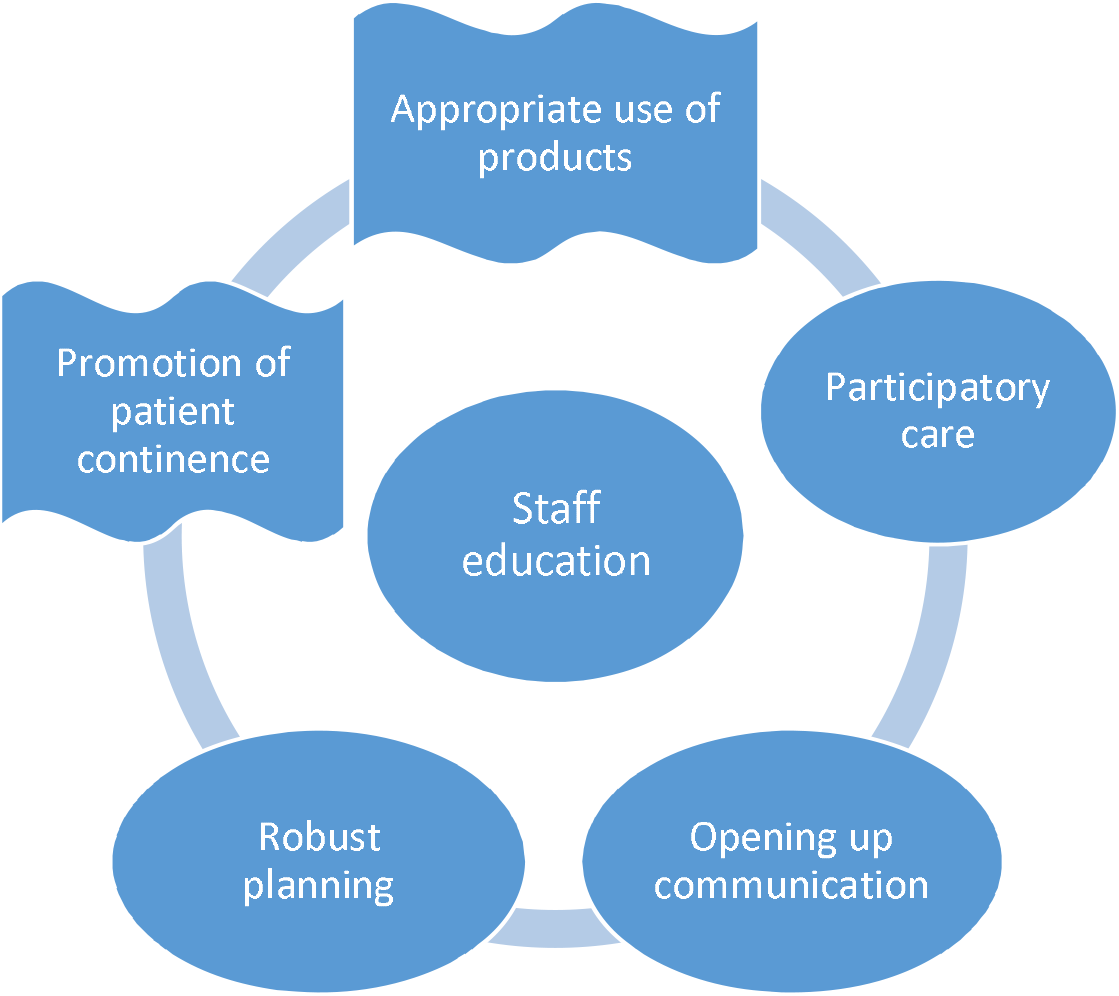
Suggested improvements to continence care practice and related outcomes.

#### Participatory care

Interviewees talked of wishing to enable greater patient independence and management of their continence, thereby helping mitigate *“institutionalised”* and *“dependent”* behaviour. This, they said, would best be achieved through a facilitative approach based on participatory care, both in respect of product use and mobilising to use the toilet:

> *I think we are all very well meaning as a profession but I do think we put an onus on people being dependant on us. You know, “it’s OK, I will come and wash you my lovely”. No! Put a bowl there, let them do it themselves! Get them walking out to the toilet, get a commode by the side of them, do what it takes… we need to look at that and say, “right, we are managing your care but I need you to be participating in your care and increase your independence so we can [better] meet your needs”. [A09, senior staff nurse]*.

#### Opening up communication

Participants put forward the likely benefit of opening up, or broadening, their communication with, or about, patients with incontinence. A consultant geriatrician, admitting that faecal incontinence is *“a bit under-recognised”*, observed that *“we only uncover things when we ask about them [and] we are less good asking about bowel problems than we are about bladder problems”* [C24]. More generally, interviewees suggested that talking in a matter-of-fact way, with patients about incontinence would help overcome patients’ reluctance to do so. Furthermore, attentive listening and opening up a dialogue so that patients, and, where appropriate, family members, were better able to express their views would also allow staff to fine tune interventions to meet the individual’s needs:

> *Some feedback from patients about how we can make them feel a bit more comfortable being incontinent would be good. Because sometimes you can go over to somebody and say its fine, don’t worry. It would maybe nice to know how they would want us to approach it as opposed to [us] just batting it off. [A08, senior staff nurse]*

#### Robust planning

Participants told us that continence care plans, where they existed, provided a useful guide *“so that everyone knows what the situation is and how it should be managed”* [C25, physiotherapist]. Interviewees added that to be effective, however, continence care planning had to be robust in terms of routine updating and forward thinking. Routine consideration of continence care needs would more likely take place, according to one consultant geriatrician, within an integrated appraisal system:

> *The thing that we have lost is proper old fashioned MDTs*… *now you have a daily ward round where you catch up with everyone but continence [is missed] when you are nipping round quite quickly, whereas in the past I would do MDTs where we would routinely talk about skin care, continence, nutrition. We don’t do that now. So having something where it was routinely discussed for everyone, that would help. [C24, consultant geriatrician]*.

A consultant physician also advocated a higher profile for continence care within *“intentional rounding”* [B46]. This sharper focus on continence care would, according to a staff nurse, facilitate forward planning and preventative work, with the corollary *“I think we can then avoid the need for catheters”*. [A09, senior staff nurse].

Participants also wished to see more proactive planning in regard to patient discharge and associated community liaison. Particular emphasis was put on improving referral procedures for district nursing regarding catheter care, in order to mitigate delays that were said to arise when discharge letters to GPs were not promptly acted on and to enable a better *“flow”* of information to district nurses [B49].

#### Staff training

The majority of participants told us they had received no *“specific”, “formal”* training in continence care but had learned about it *“on the job”, “talking to colleagues”*, or *“just doing it”*. This was a particular concern in regard to nursing and health care assistants, given that they often worked with *“high continence needs but haven’t really had much training in it”* and so may be unaware, for example, of *“the link between skin [integrity] and continence”* [C23]. Doctors also conceded *“we get very little [continence care] training… it’s not high enough on our curriculum”* [C24], and, *“if we did have [continence care] training we might be able to stop these situations when we have forgotten to TWOC the patient”* [C29].

Participants believed continence care education would potentially have a pivotal impact on practice, improving staff awareness of relevant information, advice and best practice guidelines. Most importantly in the opinion of many participants, improved recourse to relevant education would help ensure proper, safe use of continence care products. Specific updates on incontinence pads would help provide “awareness of why we are putting a pad *on”* [C27] as well as guidance on *“how to make sure [staff] have got the right size pad”* [C29] and *“how often you change pads… we don’t know”* [B42]. As regards catheters, a therapy technician suggested staff would benefit from *“teaching around… weighing up the effects of long-term catheter and short-term catheter [use]”* [A03]. Education would also help staff *“get rid of”* incontinence sheets, which were in some cases still used inappropriately with incontinent patients and sometimes *“causes their skin to break down” [*A01].

## DISCUSSION

This study has drawn attention to incontinence as a demanding condition frequently encountered in the hospital setting, often accompanied by multimorbidity and exacerbated by poor continence management.[46,47] Findings help explain the persistence of certain barriers preventing appropriate continence care for older patients in hospital, as well as key opportunities for improvement in this setting. The significance and implications of our study findings are discussed in respect of the most prominent of these barriers and opportunities, together with the pivotal themes of assessment, communication, patient independence and staff training.

Previous studies have highlighted the importance of assessment as a benchmark of appropriate continence care.[9,18] The hospital practitioners we interviewed were certainly aware that ongoing assessment of patients with incontinence increases practitioners’ understanding of the condition as well as the efficacy of particular treatment interventions. Practitioners were also mindful of proactive treatment strategies, arising from assessment, which promote good quality, safe, person-centred, care. Strategies include early use of the trial without catheter [TWOC] procedure, scheduled toileting and prompted voiding.[17,21,40] However, practitioners did not consistently succeed in achieving this level of evidence-based assessment and care management, partly because of resource and organisational factors that impact on staff time and limit availability of specialist support and advice.[35,36,42]

Our study indicates that practitioners knew that only when patients feel they can safely discuss their continence difficulties and wishes will they have sufficient confidence to participate actively in mobilising, toileting and other aspects of the care process known to promote continence.[31,36,,48] Furthermore, facilitative communication, we suggest, may help reduce patients’ ready agreement to the use of incontinence pads, by openly addressing issues of embarrassment and reluctance to bother busy staff. In this context, future research could usefully investigate practitioner awareness of the potential for sensitive conversations to improve continence care. [49] Other studies have found that good communication is also a key component of well integrated continence care within services and between hospital and relevant community agencies, although the quality of such communication is variable.[32,37]

When practitioners spoke of wishing to help patients regain continence and return to their normal level of independence, they were, to some extent, trying to offset the patient’s likely institutionalisation in the hospital environment and its corollary, greater patient dependency or deferment of independence.[17,28,42] In this respect an important strategy is encouragement of patients to mobilise, in order to use the commode or toilet, and reduction in the use of pads and catheters. Both these aims are shown, in our study as in others, to be compromised by the interconnected obstacles of routine over-reliance on products and widespread shortages of hospital staff.[30,39] These barriers have significant consequences for patients’ health and treatment, not least in respect of gram negative bacteraemia infections, which are a national reduction priority and overwhelmingly linked with urinary tract infections and the use of urinary catheters.[50,51] Findings from this study also align with the focus in recent years on preventing pressure ulcers in the NHS. The “Stop the Pressure” campaign is now a national priority led by NHSE/I, underpinned by a national wound care strategy providing tools for NHS trusts to reduce incidence, thereby reducing harm and spiralling costs. Initial implementation achieved a 50% reduction in pressure ulcers [52] and ongoing trust accountability ensures this area of care retains this level of priority.[53] Lessons can be learned from this campaign to identify the transferable strategic framework required to raise the profile of continence care, which is the subject of a subsequent project.

Our study findings also indicate that increased training opportunities could positively address the cost implications of reactive, product-focused continence care, when compared to proactive conservative interventions, such as regular toileting, increasing mobilisation and promoting independence. This shift in approach offers the prospect of a sharper focus on the judicious use of pads and catheters to safeguard patient health and independence, which in turn requires staff to confidently initiate open discussion with patients. Beyond the hospital episode, signposting to longer term continence promotion strategies such as bladder retraining and pelvic floor muscle training could also be achieved by well-informed inpatient staff who have identified continence issues that may not have previously been discussed.[8] In line with other accounts of staff education in the context of continence care,[36,37,42,54] our data adds weight to the argument that designated continence care lead personnel, currently lacking in many hospitals, would provide the authority and oversight to help ensure this commitment to regular staff training, as well as improve staff adherence to good practice guidelines and relevant nurse proficiency standards.[55] These advances nwould aim to raise the profile of continence care as an essential aspect of good quality healthcare, requiring more than rudimentary skills.[9]

The key strengths of the study include its targeted focus on the hospital setting, an environment comparatively under researched in the context of continence health care practice. In addition to contributing insights into ways to improve continence care in hospitals, the data we gathered helps strengthen the health policy case to reduce the unnecessary use of products known to have adverse health consequences. A limitation of the study is that despite data gathering across three hospital sites, the sample size is relatively small and findings may not be generally transferable. Also, the balance of research participants was predominantly in favour of nursing staff and greater representation of medical staff and allied health practitioners would potentially have helped further refine interpretations of our research data.

## CONCLUSION

Our study set out to establish a better understanding of health carer perspectives on the barriers and enablers of good quality continence care in the hospital setting. While participants were aware of the importance of good quality continence care, they identified a number of obstacles. The study identifies the likely benefits of a more proactive and participatory engagement between practitioners and patients, achieved through effective communication and robust assessment and care planning. Such improvements are not overly complicated or costly, especially if underpinned by a commitment to provide regular training. A system-wide approach to raising the profile of this fundamental aspect of care, bringing it in line with other essential areas of patient care, will deliver tangible benefits to individuals and the NHS.

## Data Availability

Reasonable requests for data sharing will be considered.

## Acknowledgments

We would like to thank the study participants for their valuable contributions and for sharing their views so openly.

## Funding

This study was funded by University Hospitals Bristol and Weston NHS Foundation Trust, Research Capability Funding (RCF) stream.

## Competing interests

None declared

## Ethics approval

The University of the West of England Research Ethics Committee. Ethical approval given (Ref: HAS.19.07.221).

## Notes

### Competing Interest Statement

The authors have declared no competing interest.

### Funding Statement

University Hospitals Bristol NHS Foundation Trust

